# Out of Pocket Spending on Insulin under $35 Copayment Caps

**DOI:** 10.1101/2022.12.21.22283784

**Authors:** Baylee F. Bakkila, Sanjay Basu, Kasia J. Lipska

## Abstract

The high cost of insulin has made it inaccessible to many patients who need it. The Inflation Reduction Act (IRA), which was recently signed into law, limits out of pocket (OOP) spending on insulin to $35 for a 30-day supply for individuals covered by Medicare. In this study, we estimate substantial OOP savings for individuals who require insulin, finding that Medicare-insured persons are projected to save nearly one-fifth of their expenditures on insulin.

## Introduction

The high cost of insulin has made it inaccessible to many patients who need it (1,2). People with diabetes, providers, and policymakers have advocated for measures to reduce insulin costs, as list prices surge and national need for insulin increases (3). The Inflation Reduction Act (IRA), which was recently signed into law, limits out of pocket (OOP) spending on insulin to $35 for a 30-day supply for individuals covered by Medicare (4). Yet, despite the impending implementation of this policy, few studies have estimated projected savings for Medicare beneficiaries and potential savings for people with diabetes who are not currently included under the IRA (5,6).

## Methods

We estimated reductions in OOP spending on insulin under a 30-day cap of $35, using data on prescription fills from the Medical Expenditure Panel Survey (MEPS, 2017-2018). MEPS provides person- and prescription-level data on quantity, dosage, days supplied and OOP spending, with statistical weights enabling nationally-representative estimates. Any individual with at least 1 insulin fill was included. The entity that contributed the greatest dollar amount during the coverage period was designated as the primary form of insurance. Individuals who paid for insulin entirely OOP were designated as 100% self-pay.

To cap out of pocket spending at $35 for a 30-day supply, we examined spending for each prescription and its respective days supplied. We divided a prescription’s days supplied variable by 30 to standardize to a 30-day supply and multiplied by 35 to find the appropriate cap for that specific prescription. If an individual’s out of pocket spending on a prescription was greater than its calculated cap, the spending was reduced to the amount of the cap. For prescriptions missing days supplied (2,923 out of 13,235), an individual’s insulin requirement was calculated from their other insulin prescriptions. If an individual did not have any insulin prescriptions with days supplied (470 individuals out of 1,676), we imputed the dataset’s average units/day value, of 61.44.

Paired t-tests were used to calculate the difference in projected annual OOP spending (within-person) before and after implementation of the cap.

## Results

Out of 1,676 individuals who filled an insulin prescription in 2017-2018 (representing nearly 8 million people per year), 41.1% were covered by Medicare, 35.7% by private insurance, and 2.2% paid for all insulin prescriptions entirely OOP. If a $35 cap were applied to all individuals surveyed, 563 (33.6%) would see OOP savings.

Individuals with Medicare spent a median $122.67 [IQR 25.05, 406.65] annually on insulin. Under the IRA cap, individuals were projected to spend a median $102.74 [IQR 25.05, 251.86] annually (Table). On average, projected savings were $164.43 (95% CI 130.31, 198.55) per person per year, or 18.3% (95% CI 16.2%, 20.4%) less annually. Although the IRA does not cover privately insured or uninsured individuals, we estimated potential savings for these groups as well (Table).

## Limitations

This study has limitations. First, days supplied variable was missing for some insulin prescriptions, requiring imputation of dataset averages. However, where possible, we calculated an individual’s average unit requirements, which provides more personalized imputation. Second, MEPS data did not include dates of prescription fills. We applied the cap to estimated average monthly expenditures on insulin. Third, our study likely underestimates OOP savings given the pervasiveness of insulin rationing (1). Therefore, the current OOP spending we report likely underestimates potential spending if patients took their insulin as prescribed.

## Discussion

Copayment caps would result in substantial OOP savings for individuals who require insulin. Medicare-insured persons, who are included under federal legislation, will be expected to save nearly one-fifth of their OOP expenditures on insulin.

## Data Availability

All data produced in the present study are available upon reasonable request to the authors.

https://www.meps.ahrq.gov/

## Acknowledgements

SB reports funding from the National Center for Chronic Disease Prevention and Health Promotion (U18DP006526), the National Institute of Diabetes and Digestive and Kidney Diseases (P30DK092924, R01DK116852), and personal fees from the University of California San Francisco, Waymark and HealthRight 360. KL reports grants from the National Institutes Health and royalties from UpToDate where she writes/edits content. BB reports no conflicts of interest.

---

Projected Annual Out-of-Pocket (OOP) Savings on Insulin Under the $35 Monthly Cap, based on the Medical Expenditure Panel Survey (MEPS, 2017-2018).

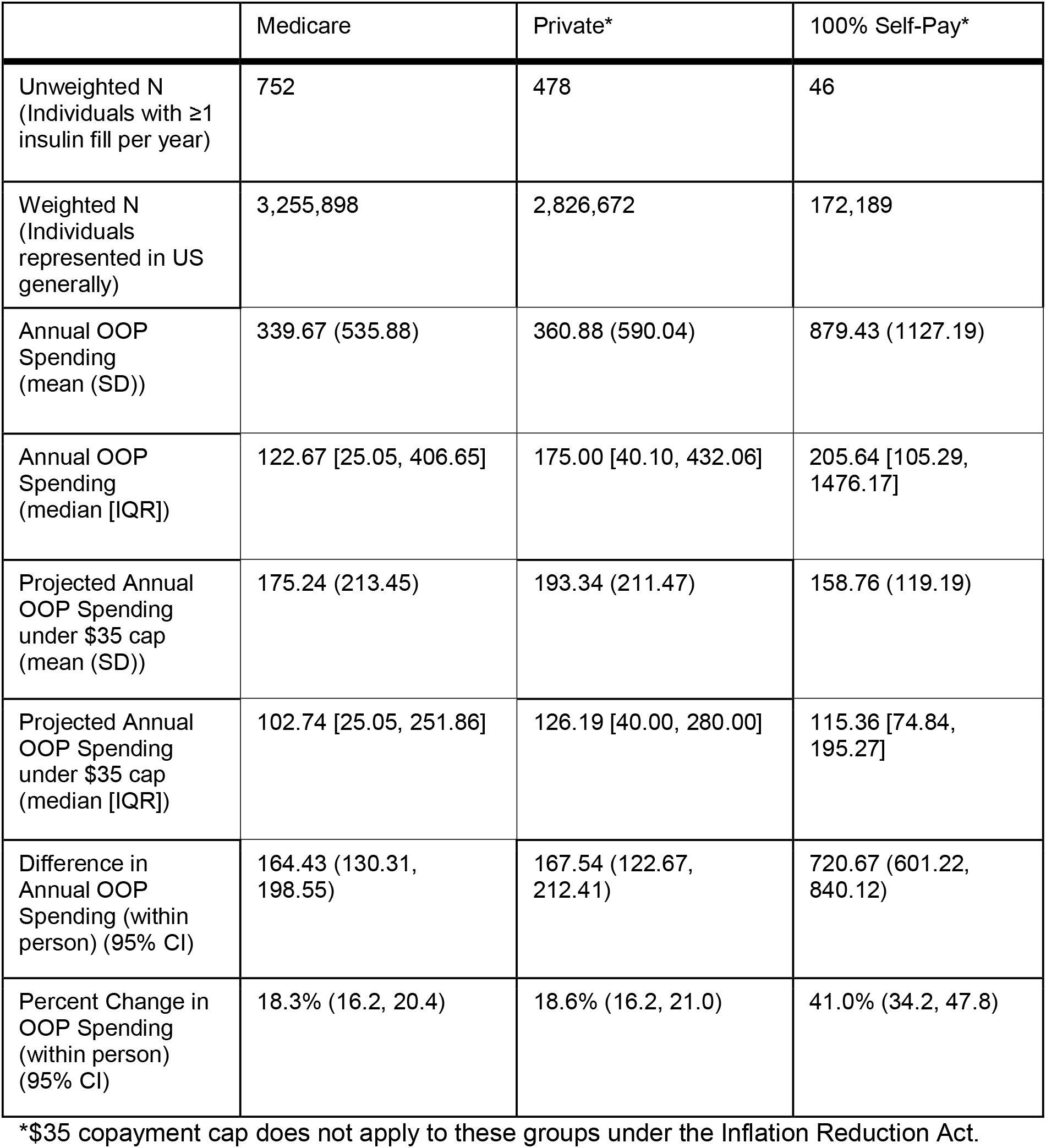

|  | Medicare | Private* | 100% Self-Pay* |
| --- | --- | --- | --- |
| Unweighted N<br>(Individuals with ≥1<br>insulin fill per year) | 752 | 478 | 46 |
| Weighted N<br>(Individuals<br>represented in US<br>generally) | 3,255,898 | 2,826,672 | 172,189 |
| Annual OOP<br>Spending<br>(mean (SD)) | 339.67 (535.88) | 360.88 (590.04) | 879.43 (1127.19) |
| Annual OOP<br>Spending<br>(median [IQR]) | 122.67 [25.05, 406.65] | 175.00 [40.10, 432.06] | 205.64 [105.29,<br>1476.17] |
| Projected Annual<br>OOP Spending<br>under \$35 cap<br>(mean (SD)) | 175.24 (213.45) | 193.34 (211.47) | 158.76 (119.19) |
| Projected Annual<br>OOP Spending<br>under \$35 cap<br>(median [IQR]) | 102.74 [25.05, 251.86] | 126.19 [40.00, 280.00] | 115.36 [74.84,<br>195.27] |
| Difference in<br>Annual OOP<br>Spending (within<br>person) (95% CI) | 164.43 (130.31,<br>198.55) | 167.54 (122.67,<br>212.41) | 720.67 (601.22,<br>840.12) |
| Percent Change in<br>OOP Spending<br>(within person)<br>(95% CI) | 18.3% (16.2, 20.4) | 18.6% (16.2, 21.0) | 41.0% (34.2, 47.8) |

\*\$35 copayment cap does not apply to these groups under the Inflation Reduction Act.

